# Assessing the clinical meaningfulness of slowing CDR-SB progression with disease-modifying therapies for Alzheimer disease

**DOI:** 10.1101/2024.07.16.24310511

**Authors:** Sarah M. Hartz, Suzanne E. Schindler, Marissa L. Streitz, Krista L. Moulder, Jessica Mozersky, Guoqiao Wang, Chengjie Xiong, John C. Morris

## Abstract

**INTRODUCTION:** For many patients and caregivers, a major goal of disease-modifying treatments (DMT) for Alzheimer disease (AD) dementia is to extend independence in instrumental and basic activities of daily living (IADLs and BADLs). The goal of this study was to estimate the effect of treatments on the time remaining independent in IADLs and BADLs.

**METHODS:** Participants at the Knight Alzheimer Disease Research Center were selected who were potentially eligible for recent DMT trials: age ≥ 60 years at baseline, clinical diagnosis of very mild or mild AD dementia (global Clinical Dementia Rating® (CDR®) score 0.5 or 1), biomarker confirmation of amyloid pathology, and at least one follow-up CDR assessment within 5 years. For IADLs, a subset of the Functional Assessment Questionnaire (FAQ) was examined that rated the degree of independence in the following: paying bills, driving, remembering medications and appointments, and preparing meals. For BADLs, the Personal Care domain of the CDR was used. Mixed-effects logistic and ordinal regression models were used to examine the relationship between CDR Sum Boxes (CDR-SB) and the individual functional outcomes and their components. The change in CDR-SB over time was estimated with linear mixed effects models.

**RESULTS:** 282 participants were followed for an average of 2.9 years (SD 1.3 years). For 50% of individuals, loss of independence in IADLs occurred at CDR-SB>4.5 and in BADLs at CDR-SB>11.5. For individuals with a baseline CDR-SB=2, treatment with lecanemab would extend independence in IADLs for 10 months (95% CI 4-18 months) and treatment with donanemab in the low/medium tau group would extend independence in IADLs by 13 months (95% CI 6-24 months).

**DISCUSSION:** Independence in ADLs can be related to CDR-SB and used to demonstrate the effect of AD treatments in extending the time of independent function, a meaningful outcome for patients and their families.

## Introduction

Alzheimer disease (AD) is a devastating neurodegenerative disorder characterized by the cerebral accumulation of amyloid and tau pathology that cause synaptic and neuronal injury leading to progressive dementia. Recently, anti-amyloid monoclonal antibodies have been shown to reduce cerebral amyloid burden with corresponding slowing of progression of AD dementia.^1, 2^ Based on these results, the United States Food and Drug Administration (FDA) approved lecanemab as a disease-modifying therapy (DMT) for early symptomatic AD in July 2023 and approved donanemab, also for early symptomatic AD, in July 2024. However, the degree to which these DMTs slow dementia progression is modest and has been deemed by some not to reach minimal clinical importance.^3^ Given that therapy with these agents is costly, burdensome, and associated with risk for amyloid-related neuroimaging abnormalities (ARIA), there has been some reluctance to initiate treatment with these drugs without a clearer demonstration of their clinical benefit.^4^

Any AD treatment must show a clinically meaningful benefit that outweighs its risks and costs.^5^ However, there is no consensus on what constitutes a “clinically meaningful” benefit for AD dementia.^6^ Although it is generally agreed that cognition and functional performance should be assessed,^7–10^ statistically significant differences in these scales in clinical trials may not always translate to a clinically meaningful effect as determined by the patient, their caregivers or family, and the treating clinician.^6^ Complicating matters is that current trials of anti-amyloid monoclonal antibodies have been restricted to persons with very mild to mild symptomatic AD, in whom functional outcomes likely differ from those with moderate or severe AD dementia.

Change in the Clinical Dementia Rating® Sum Boxes (CDR®-SB), a widely used global scale to determine the presence or absence of dementia and, when present, its severity^11, 12^ has been used as an outcome measure in Phase 3 clinical trials of these drugs. It assesses the influence of cognitive loss on the ability to conduct everyday activities by determining whether an individual has declined due to cognitive dysfunction in one or more of six domains: three cognitive (Memory, Orientation, Judgment + Problem Solving) and three functional (Community Affairs, Home + Hobbies, Personal Care). Each of the 6 CDR domains are rated as unimpaired (0) or very mildly, mildly, moderately, or severely impaired (0.5, 1, 2, and 3 respectively). Summing the scores of the individual CDR domains or “boxes” yields the CDR-SB as a continuous measure, with scores ranging from 0 (no impairment in any domain) to 18 (severe impairment in all domains). A global CDR score is then derived from the individual domain box scores.^12^ Individuals with a global CDR score of 0 are cognitively unimpaired whereas those who are global CDR 0.5 are very mildly impaired and those with global scores of 1, 2, and 3 are mildly, moderately, and severely impaired, respectively.

Patients and their families are typically most concerned with maintenance of independent function, which is highly related to safety and preservation of relationships.^13–16^ Independent function can be quantified by measuring the ability to perform accustomed activities of daily living (ADLs). Instrumental activities of daily living (IADLs) vary for each person but often include financial management (e.g., paying bills), driving a motor vehicle or arranging other means of transportation, meal preparation, and remembering to take medications and keep appointments. Basic activities of daily living (BADLs) are essential self-care tasks that individuals perform themselves such as bathing, dressing, grooming, toileting, and eating.

The ability to perform IADLs and BADLs at an individual’s accustomed level defines independence and is not only highly valued by patients and their families but also has significant financial implications. Indeed, in the U.S., caregivers of people with AD who lost independent function provided an estimated 18 billion hours of unpaid assistance in 2022, valued at $339.5 billion.^17^ In the United States, the average cost of residing in an assisted living facility is $56,068 per year, and the average cost of living in a nursing home is $112,556 for a private room and $98,534 for a shared room.^17^ For these reasons, preservation of independence is a highly meaningful outcome for patients and their families.

Although studies of clinical meaningfulness in the context of DMTs have investigated the ADLs, CDR-SB and time savings to clinically meaningful outcomes,^10, 18–20^ we sought an approach that would facilitate discussions between patients and providers regarding time savings to needing higher levels of care and DMTs. This study aimed to connect a key outcome measure used by clinical trials (CDR-SB) to outcomes that are more likely to be meaningful to patients and their families (independence in ADLs). More specifically, we estimated the number of months treatment with anti-amyloid monoclonal antibodies would be expected to prolong independence.

## Methods

### Participants

Community-living persons volunteered for participation in longitudinal studies at the Knight Alzheimer Disease Research Center (Knight ADRC) at Washington University. Both cognitively unimpaired and cognitively impaired participants were eligible for and agreed in principle to undergo amyloid positron emission tomography (PET) and lumbar puncture (LP) to obtain cerebrospinal fluid (CSF) for assays of Aβ and tau proteins, among other analytes. All participants underwent clinical and cognitive assessments using the Uniform Data Set (UDS),^21^ which includes interviews with a study partner who knows the participant well (generally the spouse or other relative) to enable scoring of the CDR and the determination of the likely etiological diagnosis of the cognitive impairment, if present, in accordance with standard criteria.^21^ For this study, we selected participants who met criteria similar to those enrolled in recent clinical trials of anti-amyloid antibodies:^1, 2^ participants had a clinical diagnosis of AD dementia in accordance with standard criteria with a global CDR score of 0.5 or 1 and confirmation of amyloid pathology as determined by CSF Aβ42/40<0.0673^22^ or amyloid PET Centiloid>20^23^ within 1 year of the clinical AD diagnosis. Additionally, all participants were aged 60 years or older at baseline and had at least one follow-up clinical assessment within 5 years of the baseline visit at which they were diagnosed with AD dementia. We used data from all assessments within this time period.

### Consent statement

Participants and study partners gave written, informed consent.

### Outcomes

The CDR-SB was used as a global measure for AD dementia progression.^11,12^ IADLs were assessed with the Functional Activities Questionnaire (FAQ)^24^ from the UDS in which the study partner rates the participant’s current ability to perform accustomed activities relative to their previous ability to do so. The rating choices are: “performs normally” (scored as 0), “has difficulty but does it by themselves” (scored as 1), “requires assistance” (scored as 2), or “dependent” (scored as 3). There is also an unscored “not applicable” category for participants who were unaccustomed to doing a particular activity. Based on studies of outcomes of interest to patients and their caregivers,^15, 16^ we selected 4 of the 10 activities in the FAQ as indicative of activities that represent independent living: 1) managing personal finances (e.g., paying bills, keeping financial records), 2) driving a motor vehicle or arranging travel outside of the home, 3) remembering to take medications and to keep appointments, and 4) preparing a balanced meal. We defined loss of independence in IADLs as a score of 2 (“requires assistance”) or 3 (“dependent”) in at least 3 of the 4 activities. For BADLs, we used the CDR Personal Care domain,^11^ with loss of independence in BADLs defined as a score of 2, indicating “requires assistance in dressing, hygiene, keeping of personal effects,” or 3, indicating “requires much help with personal care.”

### Statistical Modeling

All statistical analyses used SAS 9.4^25^ and/or R version 4.4.0.^26^ To model CDR-SB progression over time we used the hlme function of the R lccm package,^27^ estimating a linear mixed model (without intercept) on the change in CDR-SB from baseline (time=0) as a function of time, with random effects on slopes for individual participants. In a second model, baseline CDR global score (0.5 or 1) was included as a categorical variable interacting with time.

To model the relationship between CDR-SB and each functional outcome, we selected participants without impairment in the outcome at the baseline assessment (i.e. different subsamples were used for each outcome). IADLs and BADLs were coded as dichotomous variables, and the 4 FAQ items (personal finances, driving, remembering meds/appointments, and meal preparation) were coded as ordinal variables. For each functional outcome, we fitted generalized linear mixed-effect models to CDR-SB with participant-level random effects. The glmer function of the lme4 R package^28^ was used to fit dichotomous outcomes. We ran ordinal logistic regression for the ordinal outcomes using the mixed_model function of the GLMMadaptive R package.^29^ For independent IADLs and BADLs, we used the regression models to identify the first CDR-SB where 50% of participants were predicted to be dependent (referred to as cutoffs for IADLs and BADLs).

To estimate the time (and corresponding 95% confidence intervals) to the CDR-SB values associated with loss of independence in IADLs or BADLs, we adapted a model for estimation of time savings in AD treatment trials.^30^ Specifically, we used the statistical models of the association between time (x-axis) and CDR-SB (y-axis) from our modeling and converted them to estimate CDR-SB as a function of time with corresponding confidence intervals. For clinical trial data, we used an analogous approach to estimate time to a CDR-SB cutoff for the placebo and treatment groups based on the published changes in CDR-SB.^1, 2^

## Results

### Participant characteristics

Of the 282 participants who meet the inclusion criteria, 67% had very mild dementia (CDR 0.5) and 33% had mild dementia (CDR 1) (**Table 1**). The sample overall was 88% non-Hispanic White and 10% Black or African American. Slightly more participants were men (56%). Most participants were well-educated, with an average of 15.1 years of education. The 282 participants were followed for an average of 2.9 years.

**Table 1.**
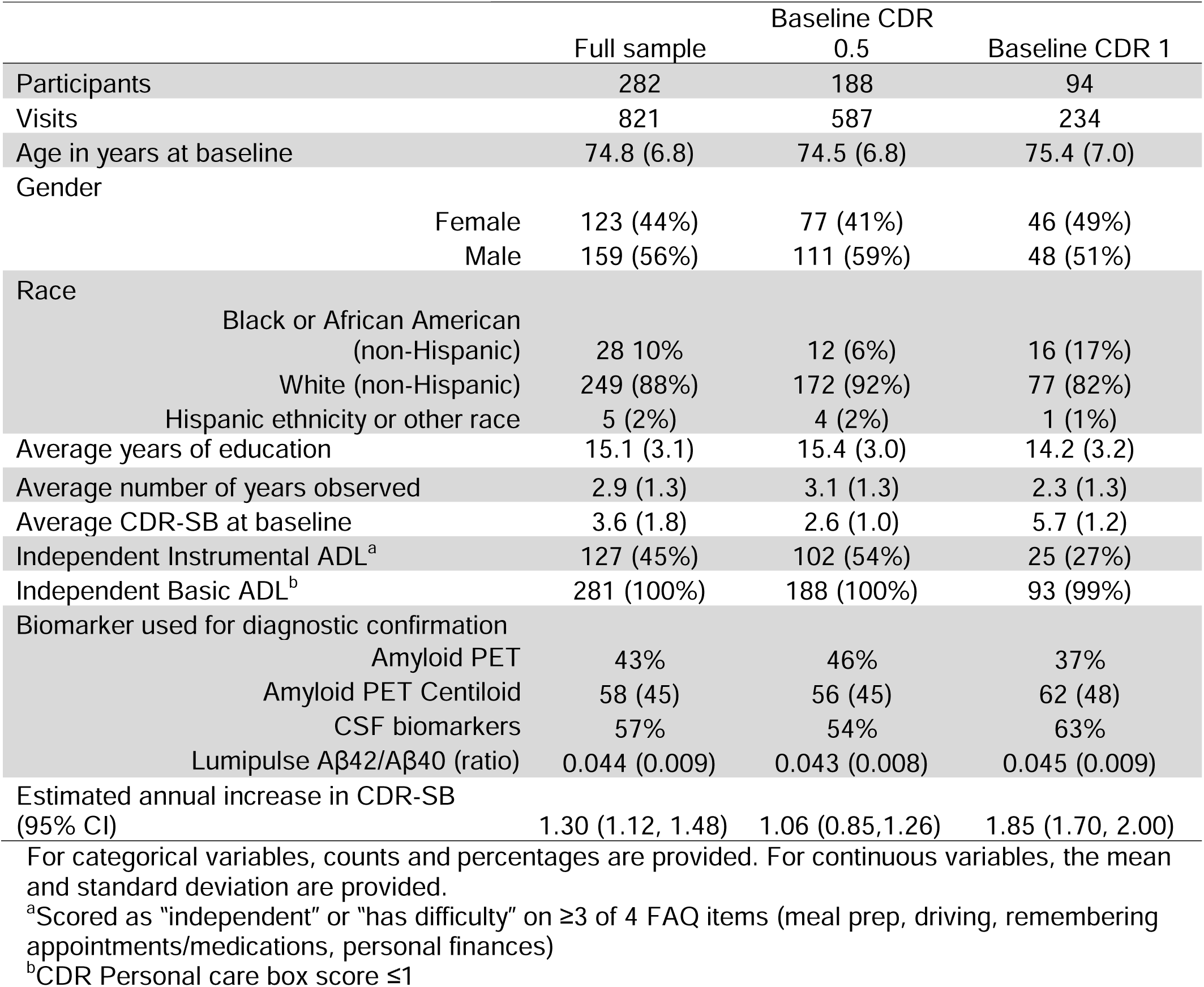
Participant characteristics.

### Relationship between IADLs, BADLs, and CDR-SB

For IADLs at baseline, most CDR 0.5 participants (95%) were independent whereas a minority of CDR 1 participants (40%) were independent (**Table 1**). Nearly all participants were independent in BADLs at baseline. We individually modeled the four individual components of IADLs (paying bills, driving, remembering medications/appointments, and meal preparation) and estimated the level of independence for each of the four items as a function of the CDR-SB score (**Figure 1A**). There were differences between the four components in the estimated CDR-SB score at which an estimated 50% of participants were dependent. For example, participants remained independent in meal preparation and remembering medication/appointments at a higher CDR-SB as compared to paying bills and driving.

**Figure 1.**
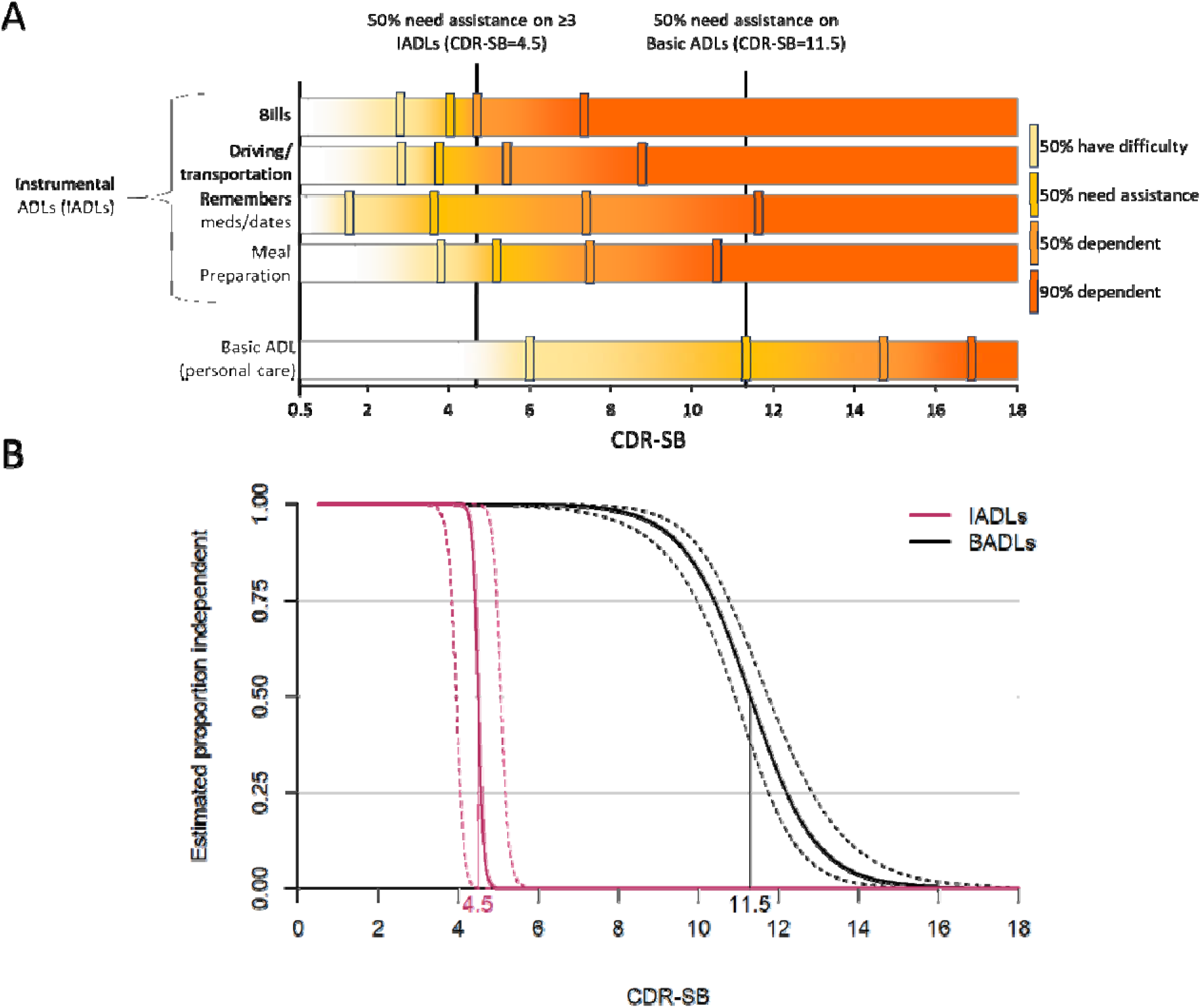
Relationship between CDR-SB and functional outcomes. Figure 1A shows the decline in components of activities of daily living (ADLs): instrumental ADLs (IADLs, personal finances, driving, remembering appointments/medications and meal preparation, with non-independence defined as “needing assistance” or “dependent” on at least 3 of these), and basic ADLs (BADLs, including dressing and personal hygiene). Figure 1B shows the probability of independence in IADLs and BADLs relative to CDR-SB. Dashed lines represent boundaries for 95% confidence intervals. The CDR-SB score is noted where an estimated 50% of participants are no longer independent in ADLs.

We next examined the relationship between CDR-SB and loss of independence in ADLs. Independence in IADLs, as defined by loss of independence with 3 or more of the four IADLs components, occurred at CDR-SB=4.5 for 50% of individuals (**Figure 1B**, based on N=352 visits from N=127 participants with independent IADLs at baseline). Loss of independence in BADLs, as defined by requiring assistance with personal care, occurred at CDR-SB=11.5 for 50% of individuals (based on N=884 visits from N=285 participants with independent BADLs at baseline). Both threshholds had very high concordance, highlighting the strong relationship between CDR and ADLs. For BADLs, 93% of participant visits scored as CDR-SB<4.5 were also scored as independent in IADLs and 87% of visits with CDR>4.5 did not have independence in IADLs. Similarly, 97% of participant visits scored as CDR-SB<11.5 were also scored as independent in BADLs and 85% of observed visits with CDR-SB>11.5 did not have independence in BADLs.

### Estimating remaining time of independence

The first step in estimating remaining time of independence in ADLs was to model progression of CDR-SB over time (change from baseline CDR-SB, **Table 1**). Overall, the average annual CDR-SB increase was 1.30 (95% CI 1.13-1.48). When modeled as a function of baseline CDR, CDR-SB increased by 1.05/year (95% CI 0.85-1.26) for individuals with a baseline CDR 0.5 and by 1.85/year (95% CI 1.70-2.00) for individuals with a baseline CDR 1. Of note, although different slopes are estimated for the different baseline CDR global values, the model assumes a linear increase in CDR-SB over time.

The modeled rate of increase in CDR-SB was then used to estimate the remaining time of independence in IADLs and BADLs. First we estimated the CDR-SB over time for each baseline CDR-SB (**Figure 2A**). Next, for each baseline CDR-SB value we estimated the time to loss of independence in IADLs (time to CDR-SB=4.5) and BADLs (time to CDR-SB=11.5), with the corresponding confidence intervals (**Figure 2B**). For example, an individual with a baseline CDR-SB=2 (with a corresponding global CDR 0.5) is estimated to have an annual increase in CDR-SB of 1.05, with a 95% CI of 0.85-1.26 (**Figure 2A**). Based on this, the expected time to loss of independence in IADLs (CDR-SB=4.5) is 29 months (95% CI 24-35 months). We made analogous computations for each baseline CDR-SB, computing remaining time of independence in IADLs for baseline CDR-SB<4.5 and remaining time of independence in BADLs for baseline CDR-SB<11.5 (**Figure 2B**). This assumes linear rates of change that are constant within global CDR 0.5, and constant within global CDR 1.

**Figure 2.**
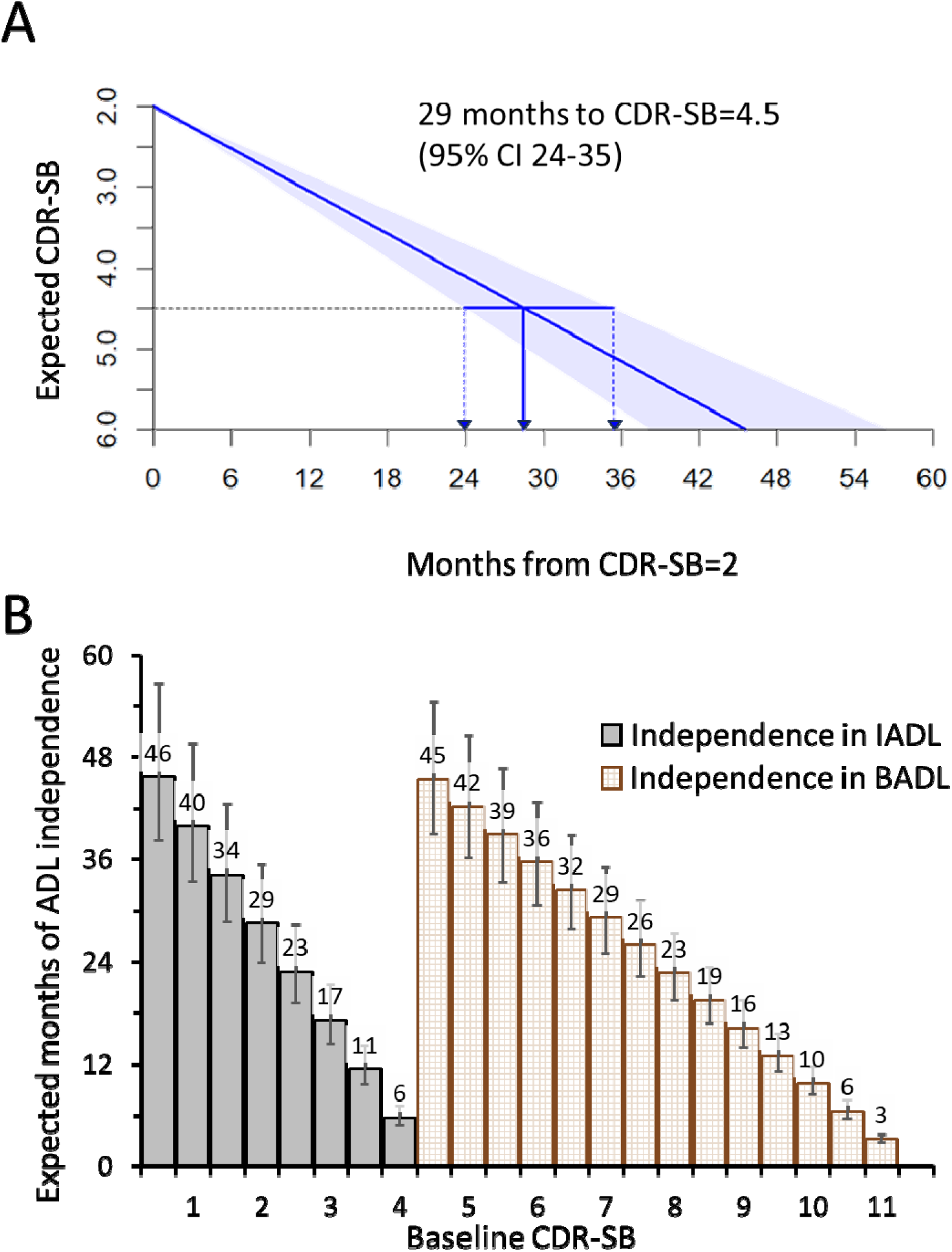
Translation of CDR-SB progression to estimated time remaining with independence in ADLs. Figure 2A illustrates the computation of expected months of independence in IADLs (CDR-SB=4.5) for baseline CDR-SB=2, using the estimated annual change in CDR-SB shown in Table 1. Figure 2B expands this calculation to baseline CDR-SB 0.5-11, showing the estimated months remaining of independence in IADLs (time to CDR-SB=4.5) for global CDR 0.5 (baseline CDR-SB 0.5-4) and independence in BADLs (time to CDR-SB=11.5) for global CDR 1 (baseline CDR-SB 4.5-11).

Finally, we estimated the additional years of independence in IADLs and BADLs associated with lecanemab treatment or donanemab treatment due to the slower rate of decline in CDR-SB (**Figure 3**). To do this, we assumed that disease progression in each arm (placebo and treatment) was linear both during and after the trial period, with a slope based on the observed effect at the end of the trial. The lecanemab trial found an average annual progression in CDR-SB for the placebo group of 1.11 (based on the published 18-month ΔCDR-SB of 1.66), with treatment decreasing this progression by 0.3 (95% CI 0.15-0.45, based on the published 18-month decrease in ΔCDR-SB by 0.45).^1^ Assuming these effects are constant over the course of treatment, those with baseline CDR-SB=2 would be expected to have an additional 10 months of independence in IADLs on lecanemab (95% CI 4-18 months, **Figure 3A**).

**Figure 3:**
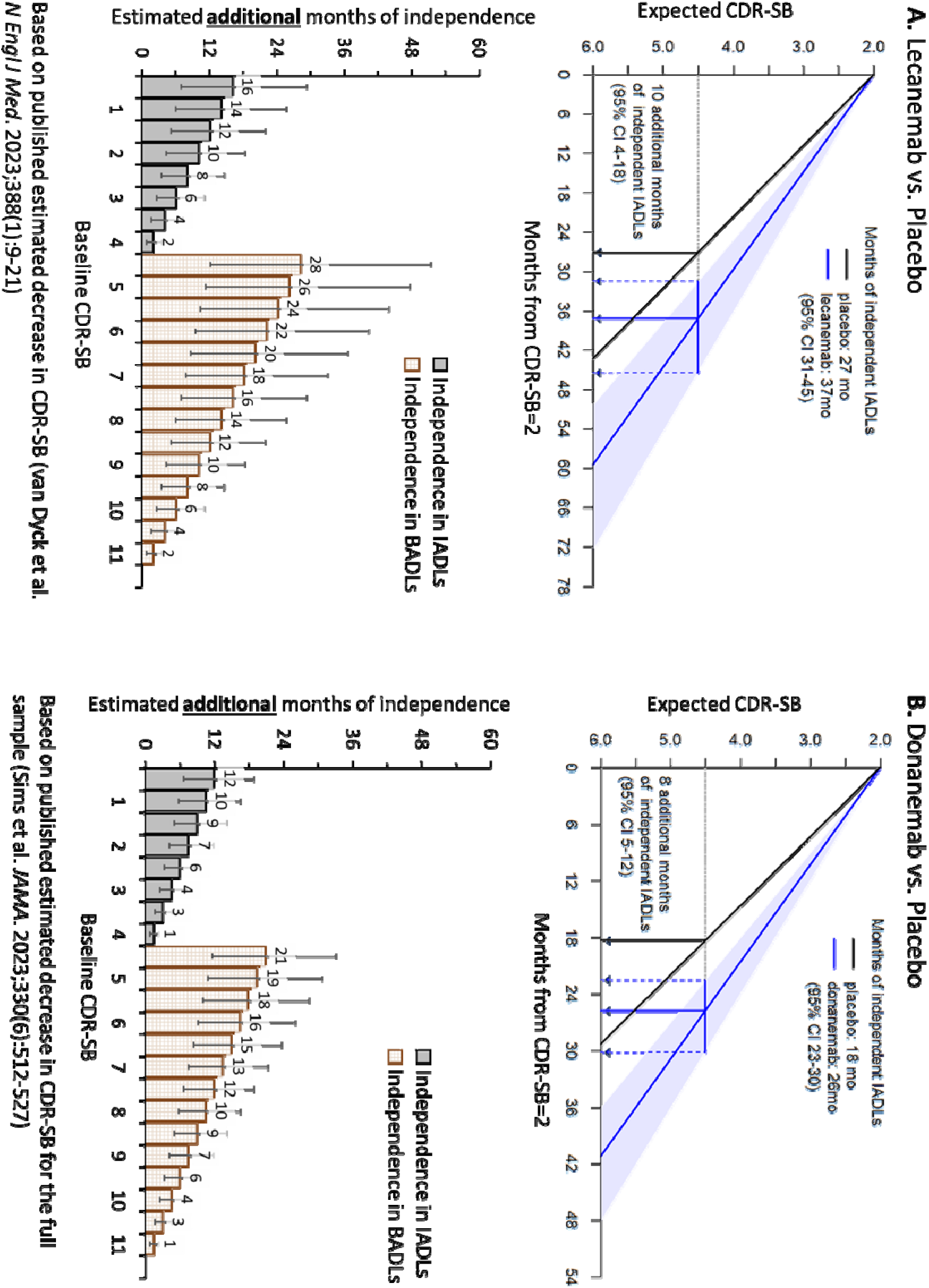
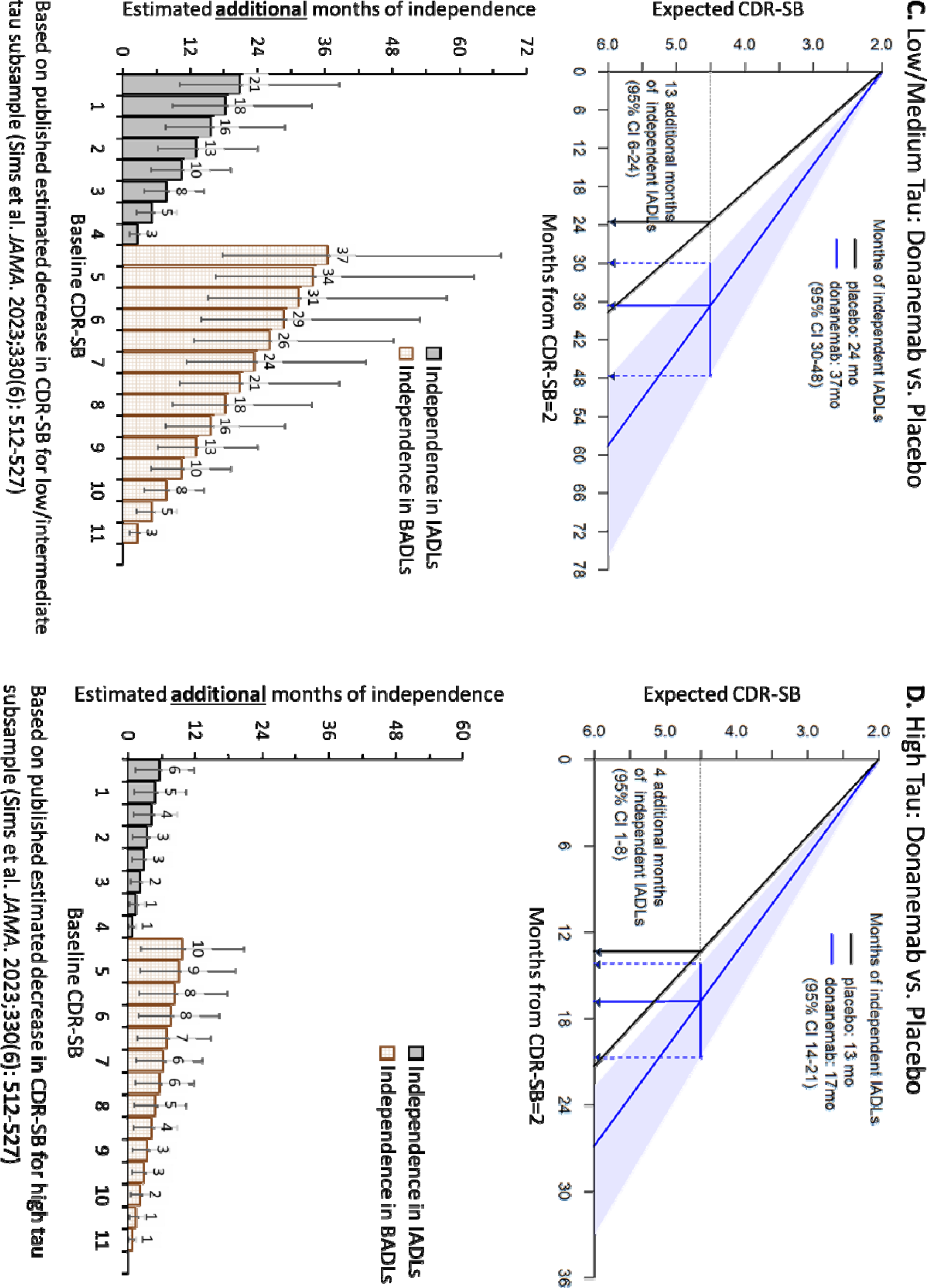
Example computation of estimated months of independence in ADLs saved by DMTs using published results. **A** shows results of the lecanemab trial. **B-D** show results of the donanemab trial: the full sample results are in **B**, and then the results are partitioned into (**C**) low/medium tau and (**D**) high tau.

Similarly, the donanemab trial found an average annual progression in CDR-SB for the placebo group of 1.65 (based on the published 76-week ΔCDR-SB of 2.42), with treatment decreasing this progression by 0.48 (95% 0.31-0.65, based on the published 76-week decrease in ΔCDR-SB of 0.7) in the combined population.^2^ The donanenab trial also reported a differential effect of treatment based on the measured tau PET levels: among those with low/medium tau PET, the placebo group had an average annual CDR-SB progression of 1.29 with donanemab decreasing progression by 0.46 (95% CI 0.27-0.65), and among those with high tau PET, the placebo group had an average annual CDR-SB progression of 2.29 with donanemab treatment decreasing progression by 0.47 (95% CI 0.14-0.82). Assuming these effects are constant over the course of treatment, those with baseline CDR-SB=2 if treated with donanemab would be expected to have an additional 8 months of independence in IADLs (95% CI 5-12 months, **Figure 3B**) in the combined population, an additional 13 months of independence in IADLs for the low/medium tau PET group (95% CI 6-24, **Figure 3C**) and an additional 4 months of independence in IADLs for the high tau PET group (95% CI 1-8 months, **Figure 3D**).

## Discussion

To better understand whether anti-amyloid treatments have a clinically meaningful benefit, we examined the relationships between the CDR-SB measure used in clinical trials and functional independence, an outcome important to patients and families. In a cohort of individuals diagnosed with early AD dementia who were confirmed to have amyloid pathology, we found that a loss of independence in IADLs occurs at CDR-SB>4.5 and in BADLs occurs at CDR-SB>11.5. We examined the rate of annual decline in CDR-SB, then estimated the time to loss of independence in IADLs (time until CDR-SB=4.5) and BADLs (time until CDR-SB=11.5). Finally, using published estimates quantifying the effects of anti-amyloid treatments on slowing decline in the CDR-SB, we estimated how long anti-amyloid treatments are expected to extend independence in IADLs and BADLs.

The time to independence in ADLs is related to the baseline functioning, and therefore additional time of independence in ADLs attributable to DMT varies. For example, an individual with a baseline CDR-SB of 2 (typically fully independent in BADLs, independent in IADLs but may have difficulty with remembering dates and medications) could expect around 10 additional months of independence in IADLs on lecanemab (95% CI 4-18 months) and 13 months of independence on donanemab (95% CI 6-24), assuming they have low/medium tau PET at baseline. In contrast, an individual with a baseline CDR-SB of 3.5 (typically fully independent in BADLs, independent in IADLs but may have difficulty with remembering dates and medications, paying bills and driving) could expect around 4 additional months of independence in IADLs on lecanemab (95% CI 2-7 months) and 5 months of independence on donanemab (95% CI 2-9 months) for those with low/medium tau PET. This interpretation of clinical meaningfulness of treatment will be particularly important for clinical decision making, allowing for personalized estimates of time to independence (based on baseline CDR-SB and, potentially, tau PET levels) that can be weighed against an individual’s risks of treatment.

Relating the CDR-SB, a global measure of AD dementia severity used in clinical trials, to independence in IADL and BADL can help patients and their families understand the benefit of treatment. The course of AD dementia from symptom onset to death generally occurs over 7-10 years,^31^ making the effects of treatment more difficult to understand when trials occur over a relatively short period (18 months). However, the slowness of AD dementia progression may magnify the potential impact of DMTs on quality of life and healthcare costs. For example, a modest slowing of disease progression as seen in the lecanemab trial could translate into an additional year of independent living for those starting with CDR-SB=1 or 9 months of independent living for those starting with CDR-SB=2. Delaying a move to an assisted living facility, with an average yearly cost of $56,068,^17^ could result in a significant savings to patients and families.

Importantly, there are methodological limitations to our study. First, to maximize the quality of our data, we limited the sample to Knight ADRC participants with biomarker-confirmed AD dementia and available longitudinal clinical data within 5 years. This restricted our sample size and may limit the generalizability of the results from the highly engaged longitudinal research cohort with biomarker-confirmed AD to a general population. Second, we assumed that CDR-SB progression is linear and that treatment effects occur uniformly during the observed study period and extend at the same linear rate beyond the length of the study. Based on other studies,^32^ cognition declines in a “waterfall” shape, suggesting that early reduction of decline may have even greater benefits at later time points. Third, we did not directly investigate the loss of function in ADLs as a function of time but rather related ADLs to CDR-SB and modeled time until a CDR-SB value associated with loss of function. While this enabled us to model treatment effects that were reported in CDR-SB, this is an indirect analysis. Future studies should explore direct associations between time and loss of ADL and confirm estimated inflection points in different samples to ensure robustness.

Despite the methodological limitations of our study, these findings can serve as a tool for understanding the relationship between the CDR-SB and functional independence. Our hope is that this modeling can be used to describe the functional implications of AD dementia progression, regardless of whether amyloid-lowering treatments are being considered. Further, this work provides an approach to better understand the clinical meaningfulness of AD treatments in terms that patients and families may find more understandable.

## Data Availability

All data produced in the present study are available upon reasonable request to the authors

## References

1. van Dyck CH, Swanson CJ, Aisen P, et al. Lecanemab in Early Alzheimer’s Disease. N Engl J Med. Jan 5 2023;388(1):9–21. doi:10.1056/NEJMoa2212948

2. Sims JR, Zimmer JA, Evans CD, et al. Donanemab in Early Symptomatic Alzheimer Disease: The TRAILBLAZER-ALZ 2 Randomized Clinical Trial. JAMA. Aug 8 2023;330(6):512–527. doi:10.1001/jama.2023.13239

3. Ebell MH, Barry HC, Baduni K, Grasso G. Clinically Important Benefits and Harms of Monoclonal Antibodies Targeting Amyloid for the Treatment of Alzheimer Disease: A Systematic Review and Meta-Analysis. The Annals of Family Medicine. 2024;22(1):50–62. doi:10.1370/afm.3050

4. Burke JF, Kerber KA, Langa KM, Albin RL, Kotagal V. Lecanemab: Looking Before We Leap. Neurology. Oct 10 2023;101(15):661–665. doi:10.1212/WNL.0000000000207505

5. Liu KY, Schneider LS, Howard R. The need to show minimum clinically important differences in Alzheimer’s disease trials. The lancet Psychiatry. Nov 2021;8(11):1013–1016. doi:10.1016/S2215-0366(21)00197-8

6. Rentz DM, Wessels AM, Annapragada AV, et al. Building clinically relevant outcomes across the Alzheimer’s disease spectrum. Alzheimer’s & Dementia: Translational Research & Clinical Interventions. 2021;7(1):e12181. 10.1002/trc2.12181

7. Siemers E, Holdridge KC, Sundell KL, Liu-Seifert H. Function and clinical meaningfulness of treatments for mild Alzheimer’s disease. *Alzheimer’s & Dementia: Diagnosis*, Assessment & Disease Monitoring. 2016;2(1):105–112. 10.1016/j.dadm.2016.02.006

8. Insel PS, Weiner M, Mackin RS, et al. Determining clinically meaningful decline in preclinical Alzheimer disease. Neurology. Jul 23 2019;93(4):e322–e333. doi:10.1212/wnl.0000000000007831

9. Wessels AM, Dennehy EB, Dowsett SA, Dickson SP, Hendrix SB. Meaningful Clinical Changes in Alzheimer Disease Measured With the iADRS and Illustrated Using the Donanemab TRAILBLAZER-ALZ Study Findings. Neurol Clin Pract. Apr 2023;13(2):e200127. doi:10.1212/cpj.0000000000200127

10. Dickson SP, Wessels AM, Dowsett SA, et al. ’Time Saved’ As a Demonstration of Clinical Meaningfulness and Illustrated Using the Donanemab TRAILBLAZER-ALZ Study Findings. J Prev Alzheimers Dis. 2023;10(3):595–599. doi:10.14283/jpad.2023.50

11. Morris JC. The Clinical Dementia Rating (CDR): current version and scoring rules. Neurology. Nov 1993;43(11):2412–4. doi:10.1212/wnl.43.11.2412-a

12. Morris JC, Ernesto C, Schafer K, et al. Clinical dementia rating training and reliability in multicenter studies: the Alzheimer’s Disease Cooperative Study experience. Neurology. Jun 1997;48(6):1508–10. doi:10.1212/wnl.48.6.1508

13. Watson J, Saunders S, Muniz Terrera G, et al. What matters to people with memory problems, healthy volunteers and health and social care professionals in the context of developing treatment to prevent Alzheimer’s dementia? A qualitative study. Health Expectations. 2019;22(3):504–517.

14. DiBenedetti DB, Slota C, Wronski SL, et al. Assessing what matters most to patients with or at risk for Alzheimer’s and care partners: a qualitative study evaluating symptoms, impacts, and outcomes. Alzheimers Res Ther. Jul 30 2020;12(1):90. doi:10.1186/s13195-020-00659-6

15. Hauber B, Paulsen R, Krasa HB, et al. Assessing What Matters to People Affected by Alzheimer’s Disease: A Quantitative Analysis. Neurol Ther. 2023/04/01 2023;12(2):505–527. doi:10.1007/s40120-023-00445-0

16. Tochel C, Smith M, Baldwin H, et al. What outcomes are important to patients with mild cognitive impairment or Alzheimer’s disease, their caregivers, and health-care professionals? A systematic review. *Alzheimer’s & dementia (Amsterdam*, Netherlands). Dec 2019;11:231–247. doi:10.1016/j.dadm.2018.12.003

17. 2023 Alzheimer’s disease facts and figures. Alzheimer’s & dementia : the journal of the Alzheimer’s Association. Apr 2023;19(4):1598–1695. doi:10.1002/alz.13016

18. Andrews JS, Desai U, Kirson NY, Zichlin ML, Ball DE, Matthews BR. Disease severity and minimal clinically important differences in clinical outcome assessments for Alzheimer’s disease clinical trials. Alzheimers Dement (N Y). 2019;5:354–363. doi:10.1016/j.trci.2019.06.005

19. Dubbelman MA, Verrijp M, Terwee CB, et al. Determining the Minimal Important Change of Everyday Functioning in Dementia: Pursuing Clinical Meaningfulness. Neurology. Aug 30 2022;99(9):e954–e964. doi:10.1212/wnl.0000000000200781

20. Lanctôt KL, Boada M, Tariot PN, et al. Association between clinical dementia rating and clinical outcomes in Alzheimer’s disease. Alzheimer’s & dementia (Amsterdam, Netherlands). Jan-Mar 2024;16(1):e12522. doi:10.1002/dad2.12522

21. Morris JC, Weintraub S, Chui HC, et al. The Uniform Data Set (UDS): clinical and cognitive variables and descriptive data from Alzheimer Disease Centers. Alzheimer Dis Assoc Disord. Oct-Dec 2006;20(4):210–6. doi:10.1097/01.wad.0000213865.09806.92

22. Barthelemy NR, Saef B, Li Y, et al. CSF tau phosphorylation occupancies at T217 and T205 represent improved biomarkers of amyloid and tau pathology in Alzheimer’s disease. Nat Aging. Apr 2023;3(4):391–401. doi:10.1038/s43587-023-00380-7

23. Pemberton HG, Collij LE, Heeman F, et al. Quantification of amyloid PET for future clinical use: a state-of-the-art review. Eur J Nucl Med Mol Imaging. Aug 2022;49(10):3508–3528. doi:10.1007/s00259-022-05784-y

24. Pfeffer RI, Kurosaki TT, Harrah CH, Jr., Chance JM, Filos S. Measurement of functional activities in older adults in the community. J Gerontol. May 1982;37(3):323–9. J Gerontol. doi:10.1093/geronj/37.3.323

25. SAS Institute Inc. SAS 9.4. (c) 2020;Cary, NC

26. R: A language and environment for statistical computing. Version 4.0.0. R Foundation for Statistical Computing; 2020. https://www.R-project.org/

27. Proust-Lima C, Philipps V, Liquet B. Estimation of Extended Mixed Models Using Latent Classes and Latent Processes: The R Package lcmm. Journal of Statistical Software. Jun 2017;78(2):1–56. doi:10.18637/jss.v078.i02

28. Bates D, Mächler M, Bolker B, Walker S. Fitting Linear Mixed-Effects Models Using lme4. Journal of Statistical Software. 10/07 2015;67(1):1–48. doi:10.18637/jss.v067.i01

29. GLMMadaptive: Generalized Linear Mixed Models using Adaptive Gaussian Quadrature. 2023. https://drizopoulos.github.io/GLMMadaptive/

30. Wang G, Cutter G, Oxtoby NP, et al. Statistical considerations when estimating time-saving treatment effects in Alzheimer’s disease clinical trials. Alzheimer’s & Dementia. 2024;n/a(n/a)10.1002/alz.14035

31. Vermunt L, Sikkes SAM, van den Hout A, et al. Duration of preclinical, prodromal, and dementia stages of Alzheimer’s disease in relation to age, sex, and APOE genotype. Alzheimer’s & dementia : the journal of the Alzheimer’s Association. Jul 2019;15(7):888–898. Alzheimers Dement. doi:10.1016/j.jalz.2019.04.001

32. Raket LL. Statistical Disease Progression Modeling in Alzheimer Disease. Original Research. Frontiers in Big Data. 2020-August-12 2020;3 doi:10.3389/fdata.2020.00024

